# Uptake and rollout of the World Health Organization-endorsed technologies for Tuberculosis diagnosis in Africa: A systematic review of international evidence 2007-2021

**DOI:** 10.1101/2025.02.06.25321799

**Authors:** Jean de Dieu Iragena, Esther Uwimaana, Derrick Semugenze, Kevin Komakech, Achilles Katamba, Anandi Martin, Moses Joloba, Willy Ssengooba

**Author notes:** Corresponding author (JI) and (WS).

## Abstract

**Background:** The World Health Organization (WHO) has endorsed a range of diagnostic technologies for tuberculosis (TB) over the years, and at least one new endorsement is issued every year. However, little is documented about the uptake status in the WHO African Region (WHO/FR). We assessed the status of uptake of the WHO-endorsed TB diagnostics through a systematic review.

**Methods:** We conducted a systematic review of the peer-reviewed literature in French or English using PubMed, Google Scholar, and Embase for TB diagnostics endorsed by WHO between January 2007 and December 2017. We considered an extra period until December 2021 for the uptake and roll-out of the recently endorsed technologies and considered publications from the 47 countries in the WHO/AFR. We considered uptake as the use of the test in the country at any level and roll-out of the test as it is being used in the settings in which WHO endorsed for it. A summary number of articles was kept using the Preferred Reporting Items for Systematic Reviews and Meta-Analyses diagrams (PRISMA) and the study protocol was registered with ISRCTN database. We used qualitative synthesis and quantitative data analysis by STATA version 14.0.

**Results:** We recorded 3,399 articles searched in peer-reviewed databases, then we eliminated 1683 duplicates and finally kept 1,716 articles for screening and 92 articles qualified for analysis. For the uptake, out of 47 countries, the majority of articles were for Xpert MTB/RIF (XPERT) test 22 (47%), followed by Line Probe Assay (LPA), 10 (21%), and the Mycobacteria Growth Indicator Tube (MGIT) 9 (19%). For the rollout, 11 (24%) countries and 16 (36%) had publications on Lipoarabinomannan (LAM) and XPERT test use respectively. The median years (Interquartile Range: IQR) for uptake were 6 (3.2 - 7.9), 5 (2.5 - 6.5), and 2.5 (3.2 - 7.9) for MGIT, XPERT, and LPA respectively, and lower for other tests. For the rollout, the median years for MGIT, LPA, and XPERT were 7 (3 – 7), 6 (1.6 - 9.4), and 5 (4 - 7) respectively, and lower for other tests.

**Conclusion:** Our study shows that the uptake and rollout of TB diagnostic technologies upon the endorsement by WHO among WHO AFR countries is slow. Future studies are needed to document the key factors affecting rapid uptake and rollout. Strategies to document a rapid guide to inform rapid uptake and roll-out as well as best practices are highly recommended.

## Introduction

In 2023, almost 2.5 million people fell ill with tuberculosis (TB) in the World Health Organization (WHO) African region; of those, almost 1.9 million new and relapse cases were notified (76%) leaving a gap of 24% undiagnosed, unnotified [1]. To address the End TB Strategy, the WHO has endorsed different diagnostic technologies over the years and defined the level of implementation to facilitate their uptake in tiers of laboratory systems [2]. The uptake varies across countries and depends on the complexity of the technology and minimum biosafety measures that should be implemented to reduce the risk of a laboratory-acquired infection [3]. Liquid culture systems, rapid speciation tests, and phenotypic Drug Susceptibility Testing of Second-Line Antituberculosis Drugs (pDST-SLDs) were endorsed in 2007, and 2008 respectively but with limited use at National TB Reference Laboratories where biosafety was a requirement [4, 5]. Along the same line, in 2008 and 2016, WHO respectively endorsed the use of line probe assay (LPA), the GenoTypeMTBDR*plus* for the rapid detection of *Mycobacterium Tuberculosis* Complex (*MTBC)* and simultaneously resistance to rifampicin (Rif) and isoniazid (H) for initial testing instead of phenotypic DST [6, 7] and GenoTypeMTBDR*sl* to detect resistance to Fluoroquinolones (FQs) and amikacin (Am) in patients with Rif-resistant/MDR-TB in less than 24 hours and to guide initiation of an appropriate MDR-TB treatment regimen [8]. In December 2010, WHO recommended the use of Xpert®MTB/RIF assay (referred to as Xpert, Cepheid, Sunnyvale, CA, USA) to represent a paradigm shift in the diagnosis of *M. tuberculosis* as well as rifampicin resistance-conferring mutations in about 2 hours, directly from sputum in adults presumptive of MDR or HIV associated TB and was initially recommended for use at district and subdistrict laboratories [9].

In 2011, light-emitting diodes (LED) microscopy was endorsed by WHO which is 10% more sensitive and faster than conventional light microscopy (LM) using Ziehl-Neelsen staining [10, 11]. Besides all, the magnitude of the HIV pandemic, the growing burden and the spread of MDR-TB made microscopy irrelevant highlighting the need for investing in early detection of TB and rifampicin resistance [12].

In the same year 2011, a policy on Noncommercial culture and DST methods, such as microscopic observation of drug susceptibility (MODS), colorimetric redox indicator (CRI), and nitrate reductase assay (NRA) for screening patients at risk for MDR-TB [13].

In 2013, a new policy on Xpert assay with the addition of extrapulmonary TB was issued expanding its use on all people suspected of TB especially from nasogastric aspirate specimens in the pediatric population as an alternative to sputum [14, 15]. Later in 2017, Xpert®MTB/RIF Ultra (Ultra) was developed and recommended for use to overcome the issue of the suboptimal sensitivity of Xpert in smear-negative sputum samples [16].

In 2015, a policy on Lateral Flow Lipoarabinomannan (LAM) rapid diagnostic test (Alere Determine™ TB LAM Ag Alere Inc, Waltham, MA, USA) as a point-of-care test for the diagnosis and screening of active TB in the urine of people living with HIV has been issued [17]. Despite these recommendations and the evidence that implementation of AlereLAM reduces tuberculosis-related mortality, AlereLAM uptake has been slow due to its low sensitivity and specificity, restrictive eligibility criteria, reliance on CD4+ testing, and lack of advocacy and awareness [18, 19].

In 2016, a policy guidance was issued for TB LAMP [the Loopamp™ Mycobacterium tuberculosis complex (MTBC) detection kit, Eiken Chemical Company] for use as a rapid alternative to sputum-smear microscopy [17].

In this systematic review, we document the status of uptake, roll-out, and scale-up of the WHO-endorsed technologies for TB diagnosis in Africa. We present the frequency of countries reporting uptake and roll-out and the number of years taken to publish the uptake and roll-out of TB diagnostics after WHO endorsement.

## Methods

### Systematic review registration and reporting

The review protocol was registered with the ISRCTN database (Reg. no.: ISRCTN24711056) and designed following the PRISMA 2020 Statement [20]

### Search strategy

With articles published in French or English, through an electronic and manual search, using PubMed, Google Scholar, and Embase, we conducted a systematic review of the peer-reviewed literature on TB diagnostics (in humans). The diagnostics must have been endorsed by WHO between January 2007 and December 2017 and published until December 2021 in 47 countries in the WHO African region [21]. For each diagnostic technology, we searched published articles per country with a focus on the uptake, utilization, roll-out, implementation, adoption, and operation with a combination of the diagnostic search terms as follows: Xpert®MTB/RIF, GeneXpert, GeneXpert MTB/RIF, Xpert, Ultra, NAAT, nucleic acid amplification test, CBNAAT, Cartridge Based nucleic acid amplification test; LAM, Urine LAM, LF-LAM, TB-LAM, Lateral Flow LAM, Urine antigen, Lateral flow urine lipoarabinomannan, FujiLAM, AlereLAM; LAMP, TB LAMP, TB-LAMP, Eiken, loop-mediated isothermal amplification, Loopamp MTBC; LED, Light-emitting diode fluorescent microscopy, auramine staining, LED-fluorescence microscopy, FluoLED; LPA, MTBDRplus, Genotype MTBDRplus, Genotype MTBDRsl Genotyping drug susceptibility testing, Hain, line probe assay, rpoB and katG genes, Hain Lifescience, Nipro; MGIT, BACTEC MGIT 960, BACTEC 960, Mycobacterium Growth Indicator Tube, liquid culture; MODS, Microscopic Observation Drug Susceptibility, nitrate reductase assay, NRA, colorimetric redox indicator, and CRI.

All retrieved articles were downloaded from the search database and uploaded into Rayyan systematic review software [22]. We used the “blind on” mode of the Rayyan software for screening. Four reviewers (JI, EU, KK, and DS) independently screened titles, abstracts, and full text in the “blind on” mode using predefined inclusion and exclusion criteria. A disagreement between reviewers was resolved through discussion by taking into account the level of scoring (included versus excluded) between reviewers to determine the final articles eligible for full-text reading. In the case of a persistent disagreement, a fifth reviewer (WS) was consulted as a tie-breaker to reach a consensus. Eligible articles were agreed upon by all reviewers and included in the analysis. At each stage of screening, the number of excluded articles was recorded together with the reasons. All reviewers regularly met to determine if their approach was consistent with the objective. The PRISMA flow chart for systematic review was used to document the study selection process (**Figure S1**.).

The uptake refers to the action of taking up or making use of TB diagnostic technologies upon their availabilities following the endorsement and recommendations for use by the WHO and this showed articles talking about tests that were implemented at the central level versus those implemented at the intermediate (or regional) and peripheral levels of the TB laboratory network. We defined the roll-out of the test as it being used in the settings in which it was endorsed for by WHO. i.e. central, intermediate or peripheral (**Figure *1***).

**Figure 1.**
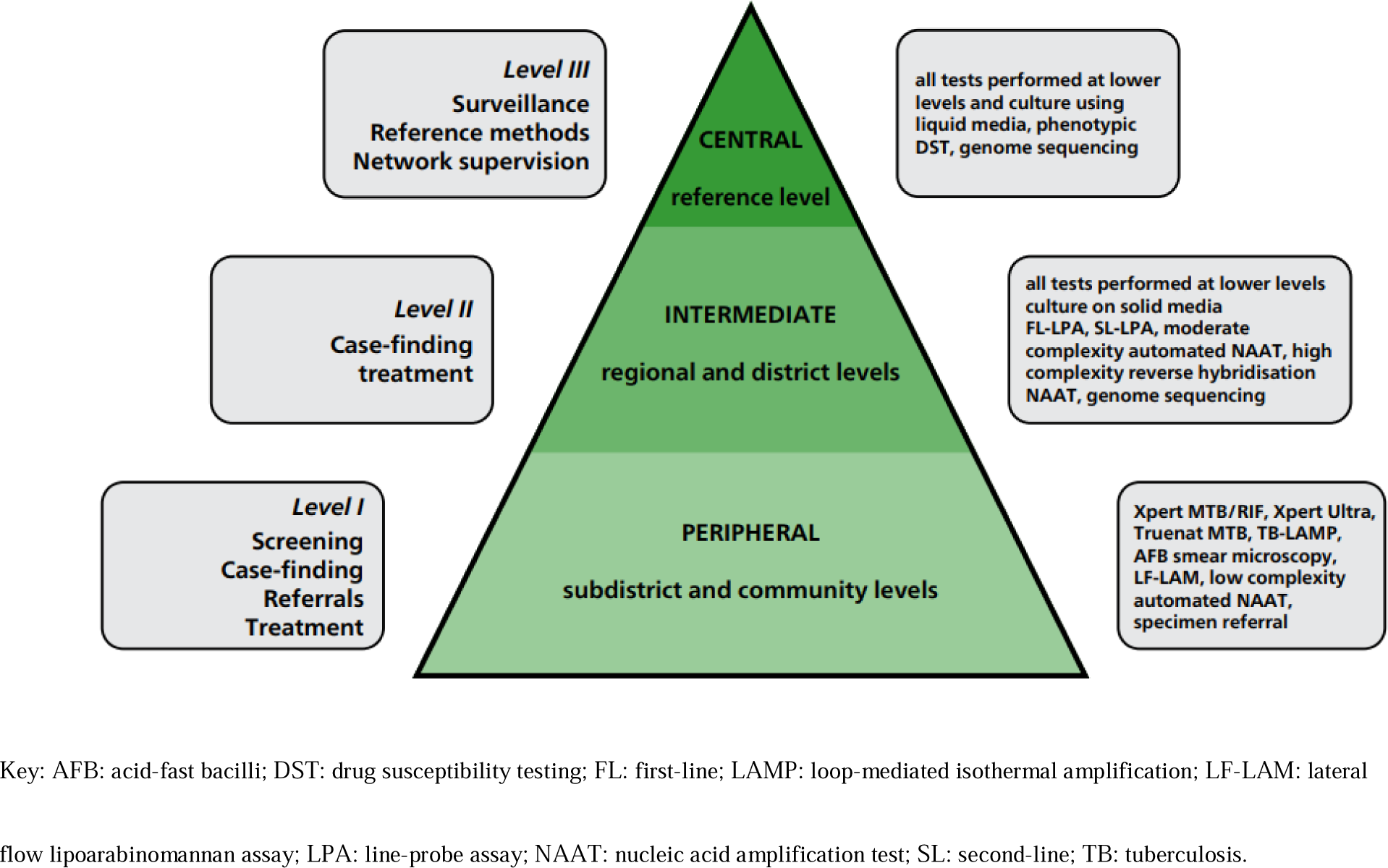
Organization of a TB diagnostic network [23].

### Eligibility Criteria

#### Inclusion criteria

We included publications on implementation at central, regional, and peripheral levels. Studies in research labs, routine-based labs, communities and research studies on the uptake and utilization of TB tests at the country level for which results were used for patient care was considered as roll-out (decentralization).

### Exclusion criteria

We excluded articles for studies conducted outside the 47 WHO/AFR countries, those without a clear African setting or specific WHO-endorsed TB test name, non-TB-specific studies, studies not referring to uptake, rollout, and implementation, studies with TB tests endorsed before 2007 or after 2017 and implemented before 2007 or after 2021. We also excluded irrelevant studies, duplicates, studies with no full text, study protocols, case reports, poster presentations, theses, studies missing years and levels of implementation, and studies with tests not yet endorsed by WHO.

### Data extraction, synthesis and Analysis

We used a standard data extraction format prepared in Microsoft Excel (Microsoft Corp, Redmond, WA, USA). Data were extracted and described based on each article’s details, including country name, a weblink per article, year of publication, title, test name, year of implementation, screening by (title, abstract, full article), and level of implementation. We specified if the implementation occurred at central, regional, and peripheral levels including the description of the setting according to the TB diagnostic network (**Error! Reference source not found.**).

We estimated the frequency and median time for diagnostic uptake and roll-out by calculating the percentage of published papers per test per country out of 47 WHO/AFR countries and the time (in years) taken for the uptake and rollout of a test respectively expressed as median years (Interquartile Range: IQR) calculated in taking into account the year of endorsement and uptake and/or rollout of a test. STATA 14 statistical software was used.

## Results

A total of 3,399 articles were obtained from the search databases. After the elimination of 1,683 duplicates, a total of 1,716 articles (**S1 File**) were recorded and screened (**Table S1, S2**). Of 1716 articles, 1034 were excluded to remain with 682 papers for full-text screening (**Table S1, S3**). We identified in total 343 articles (**Table S1, S4**) from which 92 articles (**Table S1, S5**) including 11, 14, 9, 3, 4, 31, 13, 4, and 2 tests were retained and eligible (at least one paper per country per technology) for the systematic review analysis for MGIT, LPA, LED, MODS, NRA, XPERT, LAM, LAMP, and ULTRA tests respectively (**Table *1***).

**Table 1.**
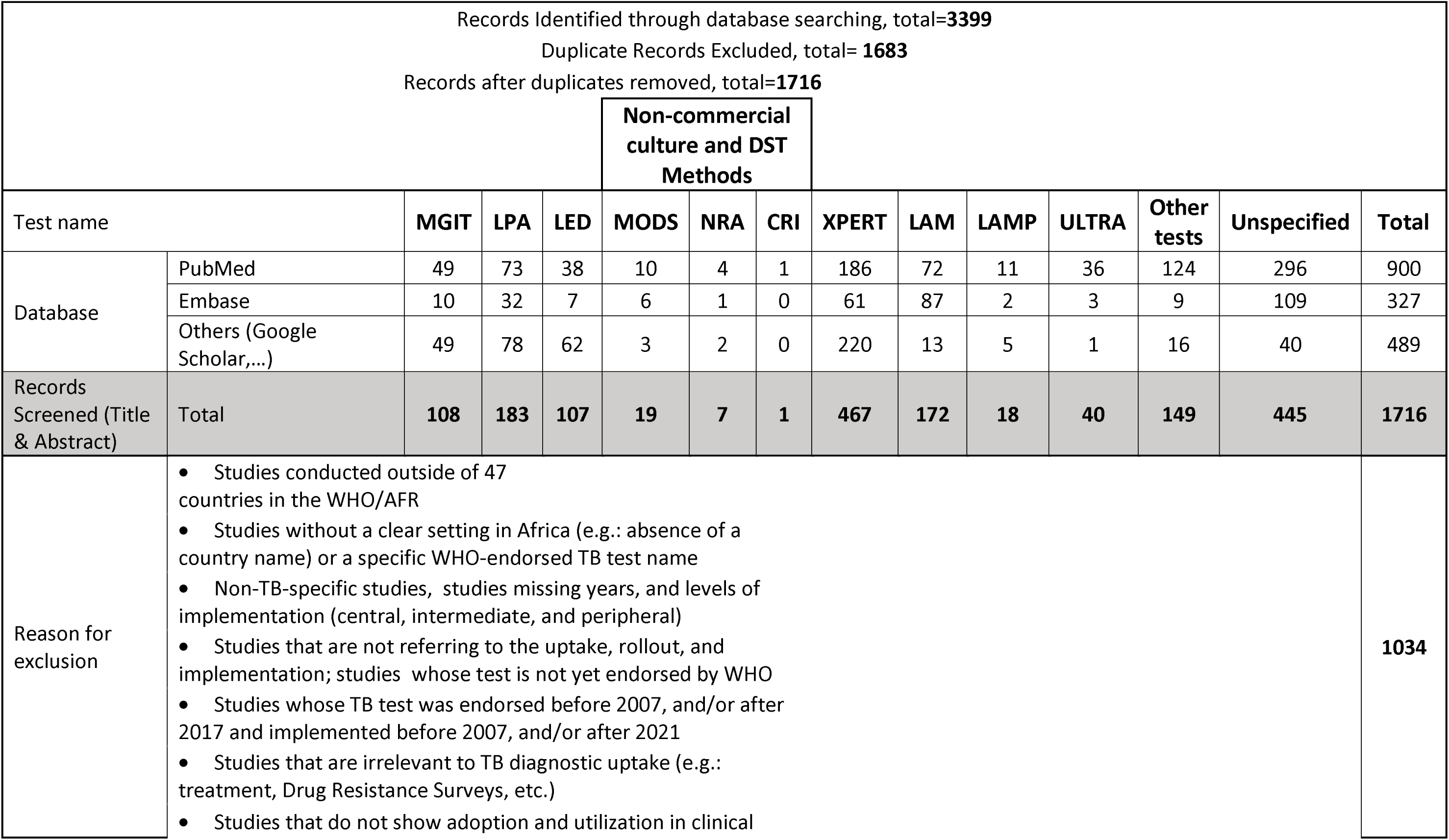
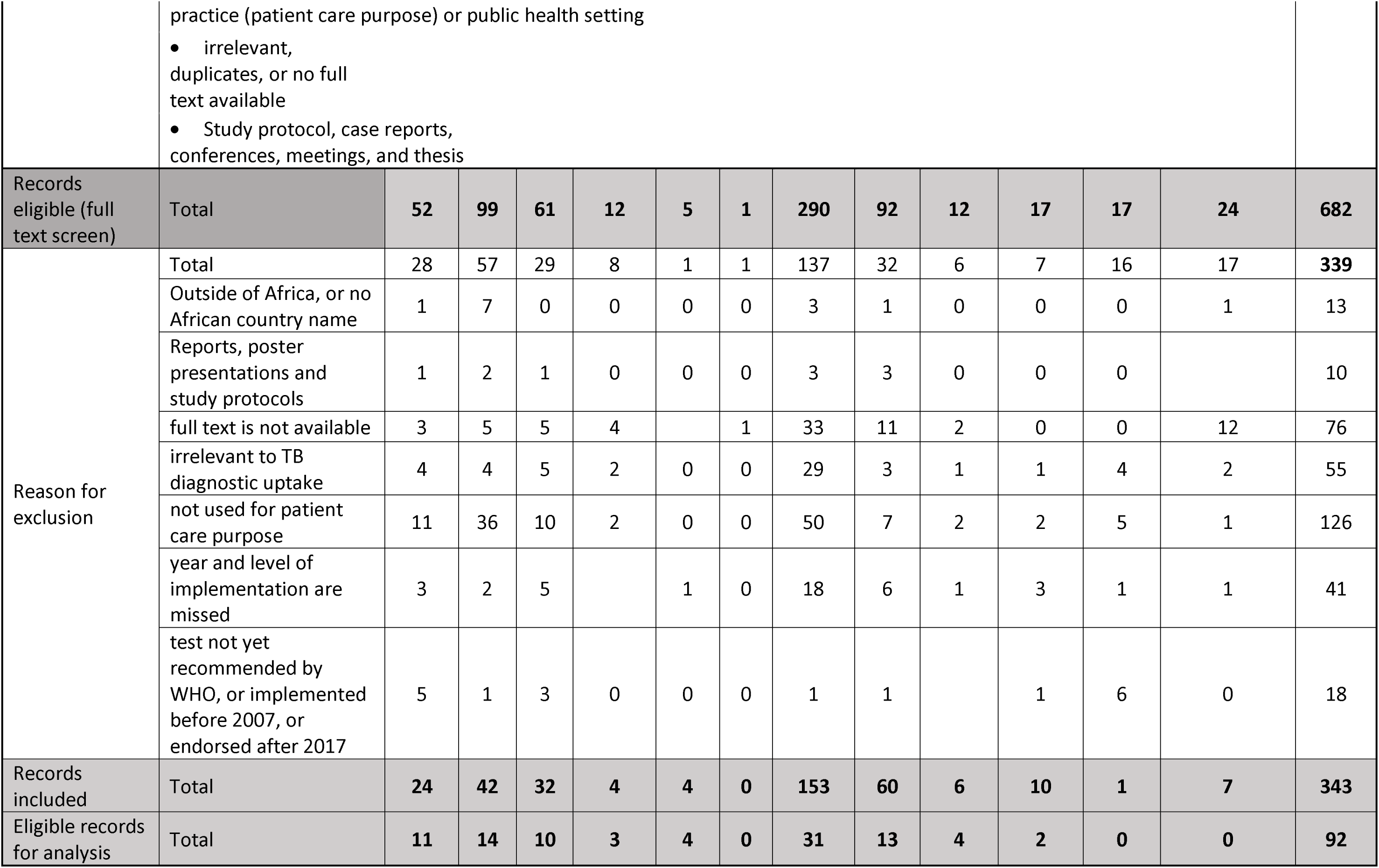
PRISMA flow diagram.

### Frequency of publications reporting the uptake and rollout of TB diagnostic tests

For the uptake, out of 47 countries, the majority of articles were for XPERT test 22 (47%), followed by LPA 10 (21%), and MGIT 9 (19%). For the rollout, 11 (24%) countries and 16 (36%) had publications on LAM and XPERT test use respectively (**Table *2***).

**Table 2.**
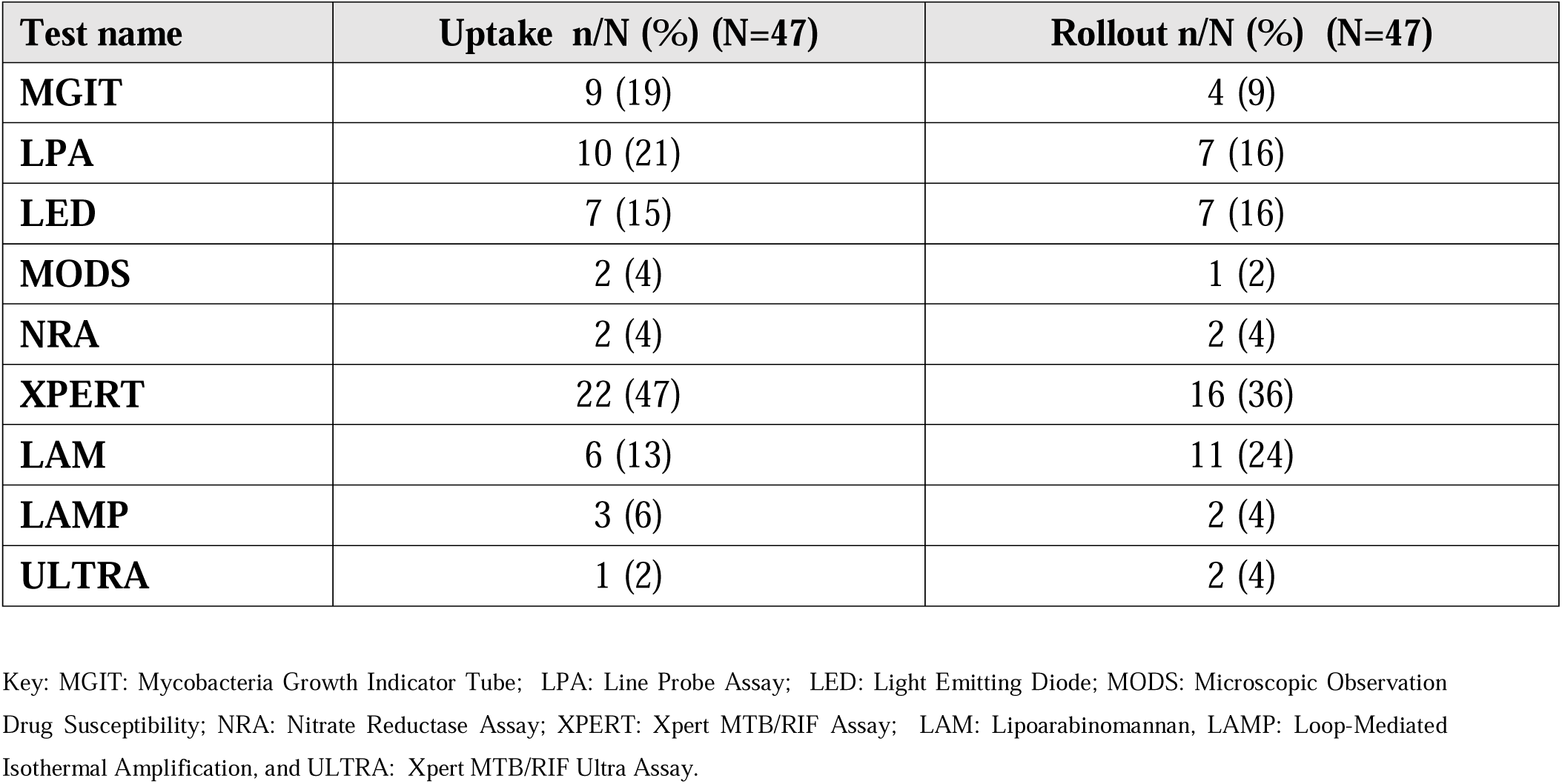
Frequency of publications reporting uptake and rollout of TB diagnostic tests.

| Test name | Uptake n/N (%) (N=47) | Rollout n/N (%) (N=47) |
| --- | --- | --- |
| MGIT | 9 (19) | 4 (9) |
| LPA | 10 (21) | 7 (16) |
| LED | 7 (15) | 7 (16) |
| MODS | 2 (4) | 1 (2) |
| NRA | 2 (4) | 2 (4) |
| XPERT | 22 (47) | 16 (36) |
| LAM | 6 (13) | 11 (24) |
| LAMP | 3 (6) | 2 (4) |
| ULTRA | 1 (2) | 2 (4) |

Key: MGIT: Mycobacteria Growth Indicator Tube; LPA: Line Probe Assay; LED: Light Emitting Diode; MODS: Microscopic Observation Drug Susceptibility; NRA: Nitrate Reductase Assay; XPERT: Xpert MTB/RIF Assay; LAM: Lipoarabinomannan, LAMP: Loop-Mediated Isothermal Amplification, and ULTRA: Xpert MTB/RIF Ultra Assay.

### Time taken by a country to report uptake and rollout of TB diagnostic tests

The median years (Interquartile Range: IQR) for uptake were 6 (3.2 - 7.9), 5 (2.5 - 6.5), and 2.5 (3.2 - 7.9) for MGIT, XPERT, and LPA respectively, and lower for other tests. For the rollout, the median years for MGIT, LPA, and XPERT were 7 (3 – 7), 6 (1.6 - 9.4), and 5 (4 - 7) respectively, and lower for other tests (**Table *3***).

**Table 3.**
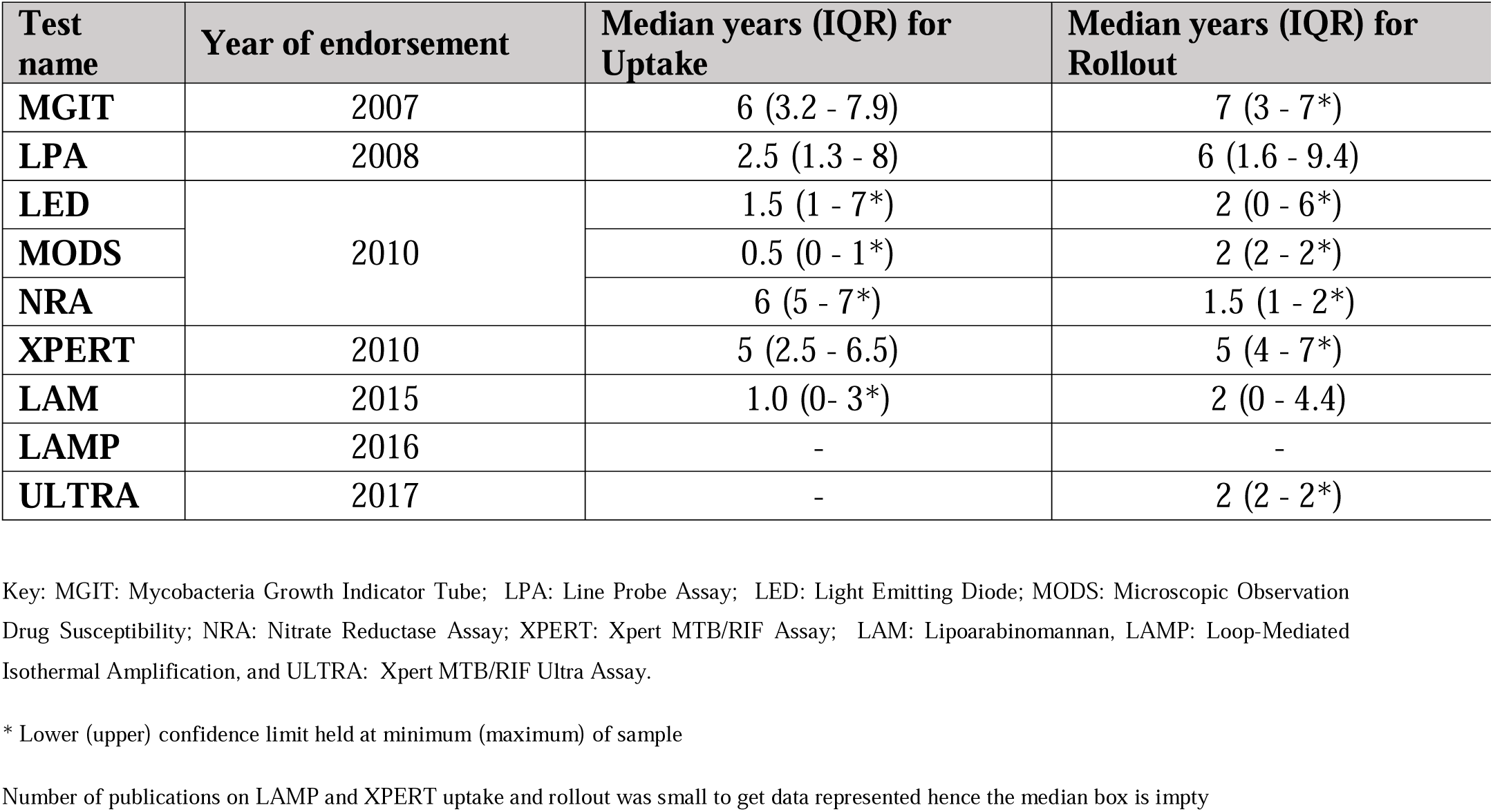
Time taken for uptake and rollout of TB diagnostic tests since endorsement by WHO.

## Discussion

Our review showed that the uptake and rollout of TB diagnostic technologies upon the endorsement by WHO is slow as may be depicted by the multiple factors affecting the uptake and roll-out of the diagnostic tests in Africa.

The review demonstrated that there is a number of publications available showing the uptake and rollout of MGIT, LPA, LED, XPERT, LAM, and LAMP progressively as they become available.

XPERT had the highest uptake and rollout across the African region, showing its use at all the levels of the laboratory network (central, intermediate, and peripheral). Our results are in line with a study on the impact and cost of scaling up GeneXpert MTB/RIF in South Africa which found a significant increase in the number of TB cases diagnosed and treated after a wide implementation of Xpert MTB/RIF across various levels of the healthcare system, demonstrating its high impact [24]. However, the delay in its uptake and rollout after its endorsement by WHO was hampered by its limitation to high-risk groups of MDR and HIV-associated TB when it was recommended by WHO in 2010 before its use was extended to extrapulmonary and children in 2017. Furthermore, its acceptability by healthcare workers as the first and most rapid molecular TB diagnostic test was slow during the early years after its endorsement.

While MGIT and LPA were primarily implemented at central (National TB Reference Laboratories) and regional levels in some countries due to biosafety requirements [25], their long median time in the uptake and rollout is in agreement with our previous findings [26] and also with what was described by Ismail et al. [27]. For instance, the uptake of LPA was 2.5 (a quick uptake as the first molecular technique in TB) while the rollout was 6 years. LPA requires specialized expertise, lengthy hands-on time, and significant laboratory infrastructure, and interpretation is not automated, limiting its utilization beyond the central level [28]. Indeed, laboratory operational delays have been noticed, including dependence on smear and culture positivity before MTBDRplus performance [29].

LAM showed significant rollout, and its penetration was more likely to have been motivated by the HIV Programmes support in uptake and roll-out as well as its easy use as a point-of-care diagnostic test without additional requirements in terms of biosafety [18]

The publications on the uptake and rollout of non-commercial culture and DST methods including MODS and NRA were very few, and this may be due to its technical complexity, cost, and the requirement for sophisticated laboratory infrastructure, as expressed by the use of these techniques has been limited in many resource-constrained settings [13].

LED microscopy took less median time than other diagnostic technologies to be implemented at all the levels. This may be due to the fact that LED required no extra infrastructural requirements and it required few days of training as personnel already had microscopy skill from the widely used smear microscopy technology. Also the fact that LED offered more diagnostic gain than conventional light microscopy among HIV-positive individuals may have provided more motivation for its rapid uptake and roll-out [30]. Although the LAMP test has good sensitivity and specificity, it is generally lower than that of the XPERT assay [31], and it does not provide a drug resistance profile. Its uptake and rollout were slow and few publications were available for analysis, hence omitted. The rollout of ULTRA was quicker (within 2 years after its endorsement), hence the uptake was not a real challenge as the GeneXpert system was already available and its use in the mindset of healthcare workers.

The slow uptake and rollout of the WHO-endorsed TB diagnostics in Africa highlights the need for improved infrastructure and training including learning from the previously successful models such as that of XPERT uptake and roll-out. It is also worth noting that countries, where the uptake and rollout were remarkable, are those that regularly participated in TB diagnostic validation studies before WHO endorsement. This may have facilitated rapid acceptability for uptake and learning lessons for rapid roll-out. Furthermore, most of the countries we found with slow uptake and roll-out are francophone, and yet most of the publishing journals are English. This may have limited the opportunity to publish articles on uptake and roll-out in such countries leading to publication bias.

Barriers and challenges linked to technical complexities, infrastructure requirements, funding and donor dependency including health care workers training and acceptance are crucial for consideration to speed up the process of uptake and rollout.

By enhancing technical assistance and capacity building for countries, increasing funding and support for infrastructure development and tailoring strategies to address specific barriers inform of implementation guide which can be used as reference in different countries may be a solution to mitigate the slow uptake and rollout.

It is essential to influence the research settings for validation and WHO Prequalification studies, which makes early awarenees of the technologies as compared to countries where these studies are usually not done. Economic situations, political commitments and instabilities may also be core factors that may influence early validation studies, uptake and roll-out in such countries with slow progress.

Our study focuses on WHO diagnostic technologies endorsed between 2007 and 2017. As publications went up to 2021 and were allowed to be included in our studies, there are other technologies endorsed between 2017 and 2021 that influenced the level of development and readiness s of some countries in improving the uptake and rollout of technologies.

In 2019, new evidence emerged justifying the use of the test in a broader group of patients, and a new urine-based test – the Fujifilm SILVAMP TB LAM, Tokyo, Japan (FujiLAM) – has been developed with 30% higher sensitivity than with AlereLAM leading WHO to issue an updated policy [32].

In 2020, a new molecular TB diagnostic tool named TrueNat which includes Truenat MTB, Truenat MTB Plus, and Truenat MTB-RIF Dx assays was developed by Molbio Diagnostics, Bangalore, India for the detection of TB and RIF-resistance from sputum sample within an hour [33].

In 2020, WHO commissioned a systematic review and recommended three classes defined by the type of technology (e.g. automated/hybridization-based NAATs), the complexity of the implementation test (e.g. low, moderate, and high - considering the requirements of infrastructure, equipment, and technical skills), and the target conditions (TB, resistance to first-line and/ or second-line drugs) [34].

In 2021, Cepheid improved the multiplexing capacity of the GeneXpert instrument (GXP) to detect a greater number of molecular targets in a single assay by upgrading the optics to a 10-color detection system (GXP10). The newly recommended Xpert MTB/XDR test detects resistance to amikacin, ethionamide, fluoroquinolones, and isoniazid, requiring the latest instrument with 10-color optics [35]

All the molecular assays endorsed so far are limited to detecting resistance to only a few first-line and second-line drugs. It is for this reason that next-generation sequencing (NGS) and bioinformatics pipelines have been developed for rapid detection of *M. tuberculosis* drug resistance, with the added advantage that sequence data could also have public health applications through understanding transmission patterns [36].

All these post-2017 endorsed technologies contributed to TB lab strengthening capacity in the African region with their specific requirements in the implementation at country levels. These rapid advances in technology brought more new and seemingly better technologies and changes in TB diagnostic policies from WHO. This may have distracted the concentration and progress toward rapid uptake of the previously endorsed technologies. However, these technological advances are giving much-needed hope in the right direction toward achieving the End TB strategy milestones and targets.

As a strength of our study, the extensive experience of the review team in TB diagnostic-related research was an asset in the conduct of systematic reviews and in collating and summarizing diverse literature in a blinded manner. Our study did a wider review coverage of 47 countries in the WHO AFR region making our findings more generalizable.

As a limitation, the uptake of diagnostic technologies as they became available has been driven by donor funding and countries may adopt the policy but the implementation or the publication is delayed or inexistent. Some countries may have also implemented it but without publication in the public domain. Therefore, some countries (low burden) were not represented and were excluded from the study. A review combining articles as well as key informant interviews or survey interviews with the countries’ implementation teams may have provided stronger findings. Other limitations may be related to potential publication bias linked to the language of publication (more papers were in English and published by English-speaking countries for instance, where French-speaking countries were few.

## Conclusion

In conclusion, this systematic review highlights the varied uptake and rollout of WHO-endorsed TB diagnostic technologies across the African region. While some technologies like Xpert MTB/RIF and LED microscopy have seen significant implementation, others face challenges due to technical complexity and infrastructure requirements. The findings underscore the need for enhanced technical assistance to capacitate countries in publishing their findings, funding, and tailored strategies to address the unique barriers faced by different countries. Future efforts should focus on comprehensive surveys to reach out to people at a country level and studies to better understand and mitigate factors influencing the uptake and rollout of TB diagnostic tools. Furthermore, designing a quick guide for technology uptake and roll-out after WHO endorsement may offer support towards technical know-how for rapid uptake and roll-out among high TB endemic countries. This may also inform the uptake and roll-out of other disease diagnostic technologies.

## Data Availability

All data produced in the present study are available upon reasonable request to the authors

## List of abbreviation

AFB: Acid-Fast Bacilli
CRI: Colorimetric Redox Indicator
DST: Drug Susceptibility Testing
FL: First-Line
LAM: Lipoarabinomannan
LAMP: Loop-Mediated Isothermal Amplification
LED: Light Emitting Diode
LF-LAM: Lateral Flow Lipoarabinomannan Assay
LPA: Line-Probe Assay
MGIT: Mycobacteria Growth Indicator Tube
MODS: Microscopic Observation Drug Susceptibility
MTBC: Mycobacterium Tuberculosis Complex
NAAT: Nucleic Acid Amplification Test
NRA: Nitrate Reductase Assay
pDST-SLDs: Phenotypic Drug Susceptibility Testing of Second-Line Antituberculosis Drugs
SL: Second-Line
TB: Tuberculosis
WHO: World Health Organization
WHO/AFR: World Health Organization African Region
XPERT: Xpert MTB/RIF Assay

## Supporting information

**S1 File.** A systematic review on TB diagnostic Upake and Rollout in Africa

**Figure S1.** PRISMA 2020 Checklist

## Acknowledgments

None.

## Authors’ contributions

JI, MJ, AK, and WS conceived the study design. JI and EU independently searched the literature in a database, designed the data extraction form, and extracted papers. DS and KK joined for the screening of papers (titles, abstract, and full text) with WS as a tie-breaker to agree on studies to be included in the qualitative analysis. JI extracted the data and wrote the initial draft. Every co-author reviewed and approved the final draft before submission.

